# Development of Predictive Risk Models for All-cause Mortality in Pulmonary Hypertension using Machine Learning

**DOI:** 10.1101/2021.01.16.21249934

**Authors:** Jiandong Zhou, Ka Hei Gabriel Wong, Sharen Lee, Tong Liu, Keith Sai Kit Leung, Kamalan Jeevaratnam, Bernard Man Yung Cheung, Ian Chi Kei Wong, Qingpeng Zhang, Gary Tse

## Abstract

**Background:** Pulmonary hypertension, a progressive lung disorder with symptoms such as breathlessness and loss of exercise capacity, is highly debilitating and has a negative impact on the quality of life. In this study, we examined whether a multi-parametric approach using machine learning can improve mortality prediction.

**Methods:** A population-based territory-wide cohort of pulmonary hypertension patients from January 1, 2000 to December 31, 2017 were retrospectively analyzed. Significant predictors of all-cause mortality were identified. Easy-to-use frailty indexes predicting primary and secondary pulmonary hypertension were derived and stratification performances of the derived scores were compared. A factorization machine model was used for the development of an accurate predictive risk model and the results were compared to multivariate logistic regression, support vector machine, random forests, and multilayer perceptron.

**Results:** The cohorts consist of 2562 patients with either primary (n=1009) or secondary (n=1553) pulmonary hypertension. Multivariate Cox regression showed that age, prior cardiovascular, respiratory and kidney diseases, hypertension, number of emergency readmissions within 28 days of discharge were all predictors of all-cause mortality. Easy-to-use frailty scores were developed from Cox regression. A factorization machine model demonstrates superior risk prediction improvements for both primary (precision: 0.90, recall: 0.89, F1-score: 0.91, AUC: 0.91) and secondary pulmonary hypertension (precision: 0.87, recall: 0.86, F1-score: 0.89, AUC: 0.88) patients.

**Conclusion:** We derived easy-to-use frailty scores predicting mortality in primary and secondary pulmonary hypertension. A machine learning model incorporating multi-modality clinical data significantly improves risk stratification performance.

## Introduction

Pulmonary hypertension is a progressive lung disorder characterized by elevated pulmonary arterial pressure, which can have different etiologies ^1^. Patients may experience symptoms like breathlessness and loss of exercise capacity that can be highly debilitating and adversely affect their quality of life. This is exacerbated by complications, such as cavitation and infection of the lungs, alveolar hemorrhage and heart failure, which can result in premature mortality ^2^. However, mortality risk differs between etiologies ^3^, and more accurate risk stratification strategies could potentially improve clinical management. To this end, several studies have developed predictive risk models based on different variables. For example, the first prognostic equation, which was based on pulmonary haemodynamics (right atrial pressure, mean pulmonary artery pressure and cardiac index at diagnosis), was derived from the National Institutes of Health (NIH) registry study of 194 patients with primary pulmonary hypertension from 32 centers ^4^. This model was applied in a contemporary cohort from the Pulmonary Hypertension Connection (PHC) registry, which showed better survival ^5^. From the Risk Evaluation and Education for Alzheimer’s Disease (REVEAL) study using a registry of 2716 patients, the model included pulmonary vascular resistance, portal hypertension, modified New York Heart Association/World Health Organization functional class IV, men >60 years of age and family history of pulmonary arterial hypertension ^6^. Recently, the Scottish composite score developed using a United Kingdom cohort of 182 pulmonary arterial hypertension patients was based on age, sex, right atrial pressure, cardiac output and 6-min walk distance ^7^.

However, to date, there have been no predictive model that could be wholly derived from data obtained from administrative databases, which would provide the opportunity to develop models based on routinely collected data to examine long-term outcomes and have the advantage over registry-based data in terms of reduced bias ^8^. Such administrative databases have been used to estimate healthcare resource utilization and costs associated with pulmonary arterial hypertension ^9^, but not for development of accurate predictive risk models. In this territory-wide cohort study, we examined the predictors of all-cause mortality, derived frailty scores predicting adverse events, and tested the hypothesis that a multi-parametric approach can improve risk prediction in patients with primary and secondary pulmonary hypertension.

## Research design and methods

### Study design

The study was approved by The Joint Chinese University of Hong Kong – New Territories East Cluster Clinical Research Ethics Committee. This population-based territory-wide cohort study included patients with a diagnosis of primary and secondary pulmonary hypertension, that were managed in hospitals under the Hong Kong Hospital Authority over the period between 1^st^ January 2000 and 31^st^ December 2017. The patients were identified from the Clinical Data Analysis and Reporting System (CDARS), a healthcare database that integrates patient information across all 43 publicly funded hospitals and their associated ambulatory and primary care facilities in Hong Kong to establish comprehensive medical records. The available information includes demographics, clinical characteristics, disease diagnoses, laboratory examinations, drug prescription details, and admission statistics. The system has been previously used by both our team and other teams in Hong Kong 10-13.

### Data extraction

Patients with primary pulmonary hypertension and secondary pulmonary hypertension were identified by their respective International Classification of Diseases Nineth Edition (ICD-9) coding of 416.0 and 416.8(3), respectively. Prior comorbidities of cardiovascular, respiratory, renal, gastrointestinal and endocrine diseases and obesity were extracted with corresponding ICD-9 diagnosis codes. Hypertension and diabetes mellitus were extracted separately. We extracted drug prescription of ten commonly prescribed drug classes: cardiac glycosides, phosphodiesterase type-3 inhibitors, thiazide diuretics, loop diuretics, potassium-sparing diuretics and, anti-arrhythmic drugs, beta blockers, vasodilative antihypertensive drugs, centrally acting antihypertensive drugs, and alpha blockers for pulmonary hypertension treatment. The mean daily dose of each drug class was reported, which is derived from multiplying the daily dose frequency against the drug dose during the study period, then averaged by the drug prescription duration. Details about the codes for identifying prior comorbidities and the specific drugs in each drug category prescribed for the study cohort are provided in the **Supplementary Tables 1 and 2**, respectively.

**Table 1.** Descriptive statistics of patients with primary and secondary pulmonary hypertension. * Mean daily drug dosage (mg/per day); * for p≤ 0.05, ** for p ≤ 0.01, *** for p≤ 0.001

|  | Primary pulmonary hypertension, n=1009 |  |  | Secondary pulmonary hypertension, n=1553 |  |  |
| --- | --- | --- | --- | --- | --- | --- |
|  | Dead; n=574 | Alive; n=435 | P value | Dead; n=574 | Alive; n=435 | P value |
|  | Median (IQR) or count (%) | Median (IQR) or count (%) |  | Median (IQR) or count (%) | Median (IQR) or count (%) |  |
| **Demographics** |  |  |  |  |  |  |
| Age | 75.29(56.22-83.82);n=574 | 48.24(16.96-68.35);n=435 | <0.0001*** | 77.11(63.08-84.89);n=575 | 48.75(6.7-67.62);n=479 | <0.0001*** |
| Sex | 186(32.40%) | 140(32.18%) | 0.3511 | 216(37.56%) | 180(37.57%) | 0.4121 |
| **Past Comorbidities** |  |  |  |  |  |  |
| Cardiovascular diseases | 480(83.62%) | 223(51.26%) | 0.3622 | 492(85.56%) | 346(72.23%) | 0.6313 |
| Respiratory diseases | 573(99.83%) | 433(99.54%) | 0.9722 | 567(98.60%) | 478(99.79%) | 0.9633 |
| Kidney diseases | 211(36.75%) | 73(16.78%) | 0.0241 | 234(40.69%) | 99(20.66%) | 0.2531 |
| Endocrine diseases | 26(4.52%) | 13(2.98%) | 0.4512 | 19(3.30%) | 18(3.75%) | 0.5231 |
| Diabetes mellitus | 94(16.37%) | 31(7.12%) | 0.0035** | 90(15.65%) | 38(7.93%) | 0.0231* |
| Hypertension | 572(99.65%) | 431(99.08%) | 0.9515 | 571(99.30%) | 478(99.79%) | 0.8924 |
| Gastrointestinal diseases | 255(44.42%) | 146(33.56%) | 0.3513 | 260(45.21%) | 161(33.61%) | 0.2531 |
| Obesity | 12(2.09%) | 14(3.21%) | 0.3412 | 10(1.73%) | 14(2.92%) | 0.1232 |
| **Prior Hospitalization** |  |  |  |  |  |  |
| No. of hospital admissions | 15.0(8.0-24.0);n=574 | 8.0(4.0-19.0);n=435 | 0.0231* | 16.0(9.0-26.5);n=575 | 12.0(6.0-22.0);n=479 | 0.1631 |
| No. of emergency readmissions | 2.0(0.0-5.0);n=574 | 0.0(0.0-2.0);n=435 | 0.0023** | 2.0(0.0-6.0);n=575 | 0.0(0.0-2.0);n=479 | 0.0032** |
| Average readmission interval, days | 198.4(102.4-355.0);n=554 | 271.7(111.8-564.4);n=396 | 0.0361* | 221.3(114.0-381.2);n=564 | 190.2(86.6-387.4);n=463 | 0.1312 |
| Cumulative length of stay | 101.0(53.5-186.0);n=574 | 36.0(15.0-109.0);n=435 | 0.1241 | 114.0(61.5-207.5);n=575 | 59.0(27.0-128.5);n=479 | <0.0001*** |
| **Drugs\*** |  |  |  |  |  |  |
| Cardiac glycosides | 108.14(62.5-241.34);n=222 | 93.75(62.50-244.98);n=63 | 0.3511 | 99.73(62.50-222.33);n=202 | 89.95(56.25-224.04);n=97 | 0.1256 |
| Thiazide diuretics | 2.50(2.17-5.00);n=152 | 3.90(2.50-19.42);n=54 | 0.2732 | 2.5(1.95-5.72);n=152 | 2.51(2.10-12.56);n=69 | 0.0451 |
| Loop diuretics | 34.35(17.64-66.15);n=497 | 27.619(10.11-58.38);n=223 | 0.0351* | 30.96(15.11-54.67);n=531 | 35.63(13.56-240.71);n=371 | 0.3521 |
| Potassium-sparing diuretics | 25.62(20.99-52.30);n=216 | 25.0(12.68-40.99);n=102 | 0.0361* | 25.36(18.00-50.02);n=207 | 14.83(8.59-28.70);n=205 | <0.0001*** |
| Anti-arrhythmic drugs | 1039.41(231.78-4227.27);n=114 | 600.0(47.70-2400.00);n=29 | <0.0001*** | 2400.0(431.52-4500.00);n=113 | 736.25(285.47-2400.00);n=50 | <0.0001*** |
| Beta adrenoceptor blockers | 34.60(18.78-84.49);n=299 | 30.2685(17.83-80.51);n=128 | 0.2631 | 30.3577(13.11-73.73);n=302 | 30.92(12.53-91.49);n=175 | 0.4511 |
| Vasodilative antihypertensives | 50.67(32.31-75.28);n=133 | 45.34(27.77-64.32);n=98 | 0.6212 | 56.66(32.82-82.13);n=115 | 53.82(36.99-60.65);n=98 | 0.3634 |
| Centrally acting antihypertensive drugs | 723.00(500.00-1749.61);n=63 | 500.0(446.96-1729.10);n=13 | <0.0001*** | 582.66(387.03-989.80);n=66 | 557.30(282.73-1572.31);n=20 | 0.2631 |
| Alpha adrenoceptor blockers | 1.70(0.89-2.93);n=93 | 1.6045(0.97-2.95);n=28 | 0.5244 | 1.58(0.82-2.69);n=119 | 2.5399(1.36-3.91);n=54 | 0.6321 |
| Endothelin receptor antagonists | 125.45(67.24-231.25);n=88 | 112.16(57.24-171.63);n=45 | 0.0125* | 135.62(48.14-179.25);n=69 | 124.17(67.24-183.66);n=55 | 0.0124* |
| Prostaglandins | 28.25(14.24-34.65);n=122 | 29.45(16.25-33.51);n=56 | 0.1252 | 29.24(19.26-35.22);n=77 | 23.55(11.45-35.25);n=69 | 0.2531 |

**Table 2.** Univariate analysis to predict mortality of patients with primary and secondary pulmonary hypertension. * for p≤ 0.05, ** for p≤ 0.01, *** for p≤ 0.001

| Variable | Primary pulmonary hypertension, n=1009 |  | Secondary pulmonary hypertension, n=1553 |  |
| --- | --- | --- | --- | --- |
|  | OR [95% CI] | P-value | OR [95% CI] | P-value |
| **Demographics** |  |  |  |  |
| Age | 1.003[1.002, 1.003] | <0.0001*** | 1.004[1.00, 1.00] | <0.0001*** |
| Sex | 1.01[0.77, 1.32] | 0.9409 | 0.99[0.78, 1.2] | 0.9965 |
| **Past Comorbidities** |  |  |  |  |
| Cardiovascular diseases | 4.85[3.6, 6.9] | <0.0001*** | 2.28[1.68, 3.10] | <0.0001*** |
| Respiratory diseases | 5.26[2.06, 8.19] | <0.0001*** | 6.15[4.02, 10.19] | <0.0001*** |
| Kidney diseases | 2.88[2.13, 3.90] | <0.0001*** | 2.63[1.99, 3.47] | <0.0001*** |
| Endocrine diseases | 1.54[0.78, 3.03] | 0.2117 | 0.88[0.45, 1.69] | 0.6906 |
| Diabetes mellitus | 2.55[1.67, 3.91] | <0.0001*** | 2.15[1.44, 3.21] | 0.0002*** |
| Hypertension | 23.12[15.24, 34.25] | <0.0001*** | 16.24[10.24, 26.44] | <0.0001*** |
| Gastrointestinal diseases | 1.58[1.22, 2.05] | 0.0005*** | 1.63[1.27, 2.10] | 0.0001*** |
| Obesity | 0.64[0.29, 1.40] | 0.2664 | 0.59[0.26, 1.34] | 0.2046 |
| **Prior Hospitalization** |  |  |  |  |
| No. of admissions | 0.998[0.99, 1.00] | 0.3089 | 0.99[0.99, 1.00] | 0.5849 |
| No. of emergency readmissions | 1.04[1.03, 1.05] | <0.0001*** | 1.04[1.03, 1.05] | <0.0001*** |
| Average readmission interval, days | 1.09[0.999, 1.20] | 0.0006*** | 1.00[0.999, 1.0] | 0.08* |
| Cumulative length of stay | 1.001[1.00, 1.002] | 0.1652 | 0.999[0.998, 1.00] | 0.1373 |
| **Drugs\*** |  |  |  |  |
| Cardiac glycosides | 3.60[2.6,5.0] | <0.0001*** | 2.10[1.6,2.8] | <0.0001*** |
| Thiazide diuretics | 2.51[1.8,3.5] | <0.0001*** | 2.21[1.6,3.0] | <0.0001*** |
| Loop diuretics | 6.01[4.5,8.2] | <0.0001*** | 3.51[2.4,5.1] | <0.0001*** |
| Potassium-sparing diuretics | 2.02[1.5,2.6] | <0.0001*** | 1.73[1.3,2.4] | <0.0001*** |
| Anti-arrhythmic drugs | 3.51[2.3,5.3] | <0.0001*** | 2.13[1.5,3.0] | <0.0001*** |
| Beta adrenoceptor blockers | 2.62[2.0,3.4] | <0.0001*** | 1.91[1.5,2.5] | <0.0001*** |
| Vasodilative antihypertensives | 1.01[0.7,1.3] | 0.8618 | 1.02[0.7,1.3] | 0.8852 |
| Centrally acting antihypertensive drugs | 4.03[2.2,7.4] | <0.0001*** | 3.01[1.8,5.0] | <0.0001*** |
| Alpha adrenoceptor blockers | 2.85[1.8,4.4] | <0.0001*** | 2.11[1.5,2.9] | <0.0001*** |
| Endothelin receptor antagonists | 0.91[0.9, 1.01] | 0.3712 | 1.34[0.9, 1.4] | 0.0561* |
| Prostaglandins | 1.01[0.91, 1.3] | 0.5311 | 1.01[0.9, 1.2] | 0.1636 |

### Primary outcome and statistical analysis

The primary outcome was all-cause mortality. Descriptive statistics were used to summarize patients’ characteristics of each primary diagnosis and mortality outcome. Continuous variables were presented as median (95% confidence interval [CI] or interquartile range [IQR]) and categorical variables were presented as count (%). The Mann-Whitney U test was used to compare continuous variables. The χ2 test with Yates’ correction was used for 2×2 contingency data, and Pearson’s χ2 test was used for contingency data for variables with more than two categories. To evaluate the significant prognostic risk factors and the effects of drug therapies associated with disease group status and primary outcomes, univariate logistic regression model was used with adjustments based on baseline characteristics. Multivariate logistic regression was conducted further to identify the important mortality factors with significant univariable predictors as input (**Figure 1**). Frailty scores without medications were derived to predict adverse events of primary and secondary pulmonary hypertension, Odds ratios (ORs) with corresponding 95% CIs and P values were reported accordingly. All significance tests were two-tailed and considered statistically significant if P values were <0.05. Data analyses were performed using RStudio software (Version: 1.1.456) and Python (Version: 3.6). Experiments were simulated on a 15-inch MacBook Pro with 2.2 GHz Intel Core i7 Processor and 16 GB RAM.

**Figure 1.**
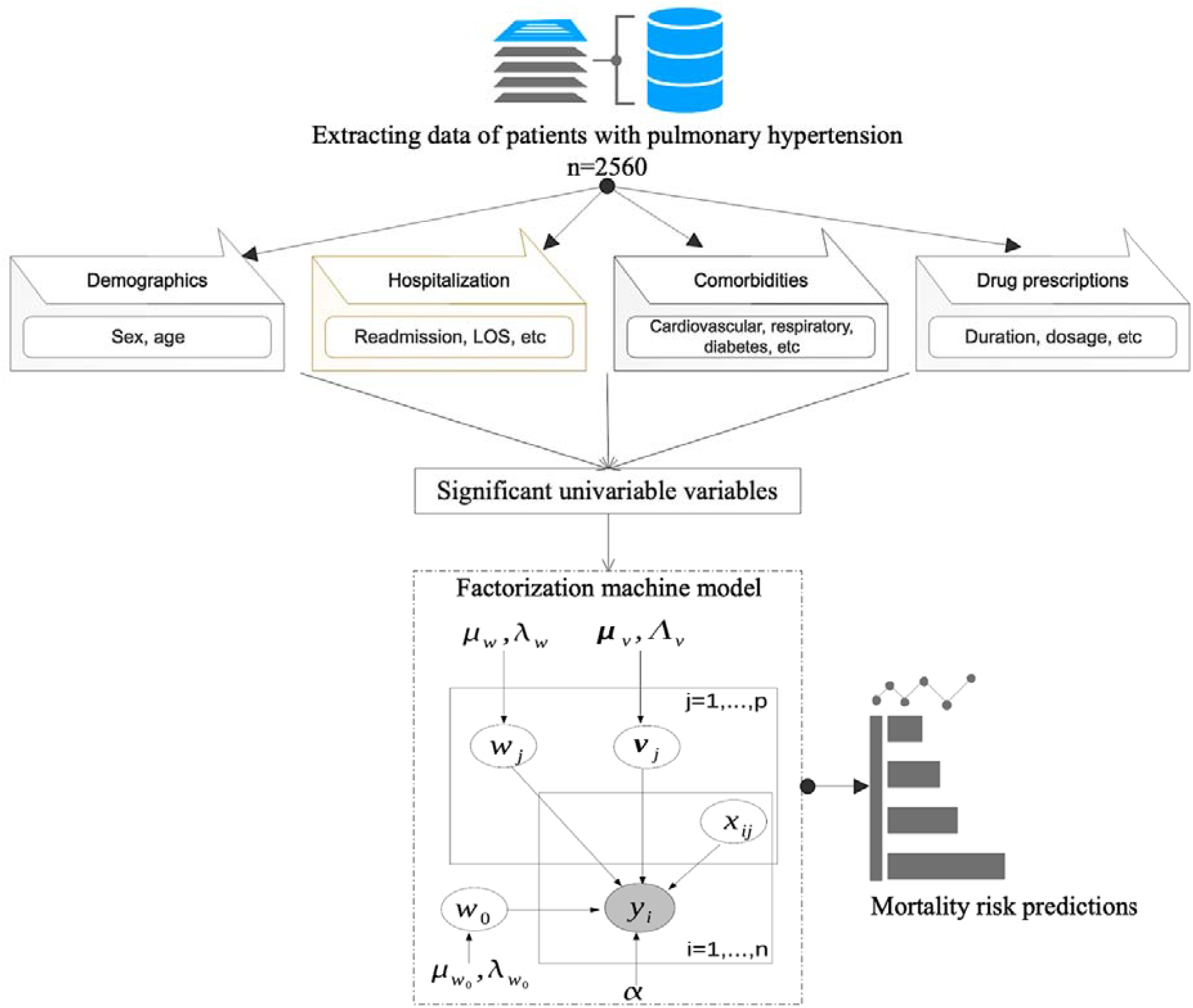
Framework of factorization machine model to predict mortality risk.

### Development of a machine learning model

In this study, we develop a factorization machine (FM) model ^14^ for pulmonary hypertension mortality risk prediction based on baseline characteristics. We observed that some categorical variables, such as comorbidities and drug prescriptions, are sparse after one-hot encoding, even leading to some missing pairs of (**x**_*j*_, **x**_*j*′_) in the training data (**x**_*j*_ and **x**_*j*′_ denotes the *j*th and *j*′ th variables). In this situation, traditional polynomial mapping approaches to capture nonlinear interactions cannot handle this data sparsity issue, while FM is able to learn interactions among variable pairs by factorizing weight matrix **W**= (**w**_1_,**w**_2_, …, **w**_|*D*|_)^T^ into **vv**^T^ where v= (***v***_l_, ***v***_2_,…, ***v***_|*D*|_) ^T^ where each row vector ***v***_***j***_ ∈**v** represents the latent vector with regard to variable **x**_*j*_. The embedding technique of the FM model can handle the hidden nonlinear and interaction patterns within the data and still demonstrate high accuracy and effectiveness when there are missing values in some features (i.e., the case of data sparsity). This motivates us to use a FM model for accurate mortality risk evaluation of patients with pulmonary hypertension based on patient’s baseline characteristics. Model performance evaluation metrics include precision, recall, F1-score and area under the receiver operating characteristic curve (AUC), and the FM model was compared to benchmark models of multivariate logistic regression, support vector machine (SVM), random forests, and multilayer perceptron (MLP).

## Results

### Baseline characteristics

The study cohort has 2562 patients, of whom 1009 and 1553 had primary and secondary pulmonary hypertension, respectively (**Table 1**). Amongst the primary pulmonary hypertension patients, 574 deaths occurred during follow-up until the end of 2019. Those who passed away were significantly older (median: 75.3, IQR:[56.2-83.8] vs. median: 48, IQR:[17.0-68.3] years old, p value <0.0001), more likely to have pre-existing diabetes, cardiovascular and renal diseases and had a shorter readmission intervals between discharges (median: 198.38 days, IQR: [95% CI: 102.37-355] vs. median 271.72 days, IQR: [111.79-564.4]; p value =0.036). However, those who survived had a similar cumulative number of hospital admissions (median: 15.0, IQR:[8.0-24.0] days vs. median: 8.0, IQR:[4.0-19.0] days; p value =0.241), number of emergency readmissions within 28 days of discharge (median: 2.0, IQR: [0.0-5.0] vs. median: 0.0 IQR: [0.0-2.0]; p value =0.6313), and cumulative length of hospital stay over the lifetime (median: 101 days, IQR:[53.5-186] vs. median: 36 days, IQR:[15-109], p value =0.1241) with those who were deceased. For secondary hypertension, 574 deaths occurred on follow-up. Those who died were significantly older and more frequently suffered from diabetes, but had a similar number of hospital admissions, number of emergency readmissions, average readmission interval but significantly longer length-of-stay. Further details on the comparisons and statistics of other baseline characteristics are shown in **Table 1**.

Drug prescription characteristics of mean daily dosages are also summarized with ten most common drug categories for both primary and secondary pulmonary hypertension patients. For primary hypertension, those who died were prescribed with a higher dosage of cardiac glycosides (median: 108.14, IQR:[62.5-241.34] mg/per day), loop diuretics (median: 34.3493, IQR: [17.6381-66.15] mg/per day), antiarrhythmics (median: 1039, IQR: [232-4227] mg/per day), beta-adrenoreceptor blockers (median: 35, IQR: [19-84] mg/per day), vasodilative antihypertensives (median: 51, IQR: [32-75] mg/per day), centrally acting antihypertensives(median: 723, IQR: [500-1750] mg/per day), alpha-adrenoceptor blocker (median: 1.7, IQR: [0.9-2.9] mg/per day) and endothelin receptor antagonists (median: 125, IQR: [67-231] mg/per day). Similar patterns were observed for patients with secondary pulmonary hypertension.

### Predictors of mortality in pulmonary hypertension

Univariate logistics regression analysis identified significant predictors of primary pulmonary hypertension mortality (**Table 2**):

1. Age (OR: 1.003, 95% CI: [1.002, 1.003], p value<0.001);
2. Prior comorbidities of cardiovascular disease (OR: 4.85, 95% CI: [3.63, 6.49], p value<0.001), respiratory disease (OR: 5.26, 95% CI: [2.06, 8.20], p value <0.001), renal disease (OR: 2.88, 95% CI: [2.13, 3.90], p value<0.001), diabetes mellitus (OR: 2.55, 95% CI: [1.67, 3.91], p value<0.001), hypertension (OR: 23.12, 95% CI: [15.24, 34.25], p value <0.0001), gastrointestinal disease (OR: 1.582, 95% CI: [1.22, 2.05], p value<0.001);
3. Healthcare utilization metrics including the number of emergency readmissions within 28 days after discharge (OR: 1.04, 95% CI: [1.03, 1.05], p value < 0.001) and average readmission interval (OR: 1.09, 95% CI: [1.00, 1.20], p value <0.001);
4. Drugs for cardiac glycosides (OR: 3.6, 95% CI: [2.6,5.0], p value<0.001), thiazide diuretics (OR: 2.5, 95% CI: [1.8,3.5], p value<0.001), loop diuretics (OR: 6.0, 95% CI: [4.5,8.2], p value<0.001), potassium-sparing diuretics (OR: 2.0, 95% CI: [1.5,2.6], p value<0.001), anti-arrhythmic drugs (OR: 3.5, 95% CI: [2.3,5.3], p value<0.001), beta adrenoceptor blockers (OR: 2.6, 95% CI: [2.0,3.4], p value<0.001), centrally antihypertensive drugs (OR: 4.0, 95% CI: [2.2,7.4], p value<0.001) and alpha adrenoceptor blockers(OR: 2.8, 95% CI: [1.8,4.4], p value<0.001).

The significant predictors of all-cause mortality for secondary pulmonary hypertension patients were largely similar to those for primary pulmonary hypertension patients (**Table 2**). Multivariate logistics regression analysis was performed by including variables with p value<0.10 (**Table 3**). After adjustment, the significant predictors were: older age (OR: 2.34, 95% CI: [1.34,3.43], p value<0.0001), prior comorbidities of cardiovascular diseases (OR: 1.79, 95% CI: [1.56,3.03], p value<0.0001), respiratory diseases (OR: 1.62, 95% CI: [1.11,2.07], p value<0.001), kidney diseases (OR: 1.21, 95% CI: [1.03, 1.45], p value=0.0001), diabetes mellitus (OR: 1.45, 95% CI: [1.02,2.16], p value=0.0003), hypertension (OR: 17.34, 95% CI: [10.43,31.27], p value<0.0001); number of emergency readmissions within 28 days discharge (OR: 1.25, 95% CI: [1.43,1.53], p value<0.0001), average readmission interval (OR: 1.13, 95% CI: [1.46,1.78], p value<0.0001). There predictors are also significant for predicting the mortality risk of the secondary pulmonary hypertension patients. A summary of the different predictors of mortality are shown in **Supplementary Table 3**. Based on the significant multivariate predictors, easy-to-use score systems were derived to predict the adverse events of primary and secondary pulmonary hypertension as shown in **Table 4**. The characteristics of patients with/without primary and secondary pulmonary hypertension using the derived score systems are further described in **Table 5**. Median risk score for identifying primary pulmonary hypertension is 11 (95% CI: [6-18], max: 27), while the median for secondary pulmonary hypertension is 12 (95% CI: [6-17], max: 28). Further, the stratification performances of score and dichotomized score systems in predicting primary and secondary pulmonary hypertension were shown in **Table 6**. Derived frailty scores demonstrated significant stratification performance in predicting primary pulmonary hypertension (OR: 1.45, 95% CI: 1.14-2.16, p value<0.0001) with cutoff 11.75 and secondary pulmonary hypertension (OR: 1.34, 95% CI: 1.2-2.01, p value<0.0001) with a cut-off value of 13.23. Dichotomized frailty scores also provide significant strengths in predicting primary pulmonary hypertension (OR: 15.32, 95% CI: 10.34-27.45, p value<0.0001) and secondary pulmonary hypertension (OR: 19.23, 95% CI: 11.88-37.21, p value<0.0001).

**Table 3.** Multivariate analysis to predict mortality of patients with pulmonary hypertension. * for p≤ 0.05, ** for p≤ 0.01, *** for p≤ 0.001

| Variable | Primary pulmonary hypertension, n=1009 |  | Secondary pulmonary hypertension, n=1553 |  |
| --- | --- | --- | --- | --- |
|  | OR [95% CI] | P-value | OR [95% CI] | P-value |
| **Demographics** |  |  |  |  |
| Age | 2.34[1.34,3.43] | <0.0001*** | 1.87[1.54,2.36] | <0.0001*** |
| **Past Comorbidities** |  |  |  |  |
| Cardiovascular diseases | 1.79[1.56,3.03] | <0.0001*** | 1.25[1.11,2.03] | <0.0001*** |
| Respiratory diseases | 1.62[1.11,2.07] | <0.0001*** | 1.43[1.12,4.23] | <0.0001*** |
| Kidney diseases | 1.21[1.03,1.45] | 0.0006*** | 1.76[1.34,2.67] | <0.0001*** |
| Diabetes mellitus | 1.45[1.02,2.16] | 0.0003*** | 1.03[1.01,1.13] | 0.0003*** |
| Hypertension | 17.34[10.43, 31.27] | <0.0001*** | 19.34[11.34,29.13] | <0.0001*** |
| Gastrointestinal diseases | 1.02[1.01,1.13] | 0.043 | 1.1[0.77,1.34] | 0.0128* |
| **Prior Hospitalization** |  |  |  |  |
| No. of emergency readmissions | 1.25[1.43,1.53] | <0.0001*** | 1.16[1.15,1.45] | 0.0003*** |
| Average readmission interval, days | 1.13[1.46,1.78] | <0.0001*** | 1.34[1.11,1.89] | <0.0001*** |

**Table 4.**
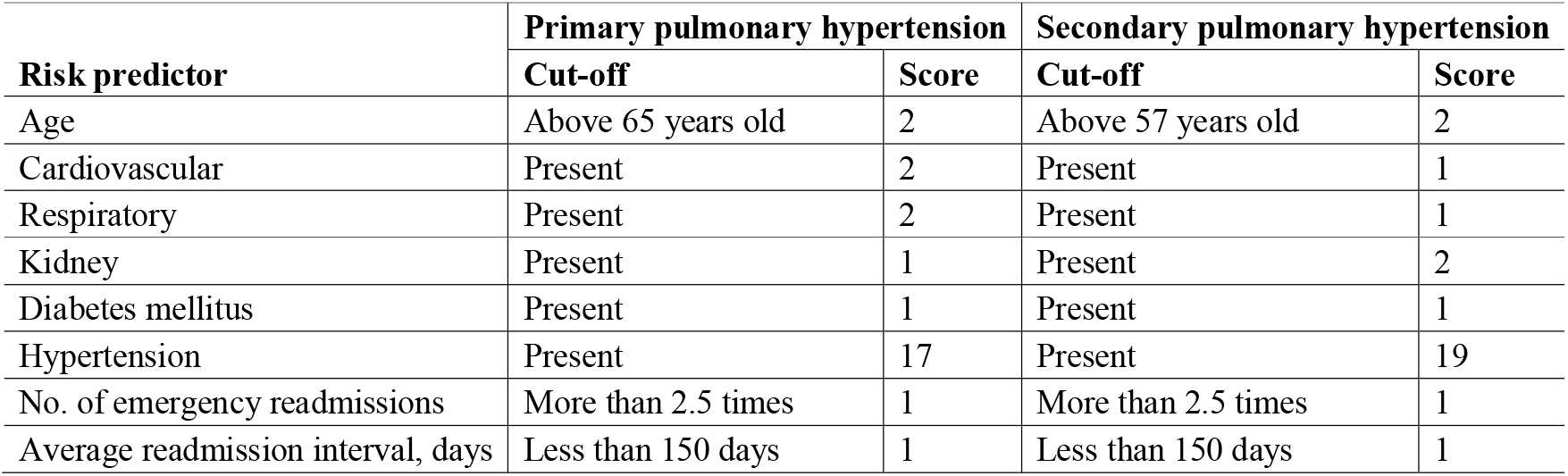
Easy-to-use score system for early prediction of primary and secondary pulmonary hypertension.

**Table 5.** Derived score characteristics of patients with/without primary and secondary pulmonary hypertension. * for p≤ 0.05, ** for p≤ 0.01, *** for p≤ 0.001

|  | No Primary Outcome<br>Median (IQR); Max | Primary Outcome<br>Median (IQR); Max | P value | No Secondary Outcome<br>Median (IQR); Max | Secondary Outcome<br>Median (IQR); Max | P value |
| --- | --- | --- | --- | --- | --- | --- |
| Derived risk score | 6(3-11);16 | 11(6-18);27 | <0.0001*** | 5(4-12);15 | 12(6-17);28 | <0.0001*** |

**Table 6.** Stratification performance of score and dichotomized score system in predicting primary and secondary pulmonary hypertension. * for p≤ 0.05, ** for p≤ 0.01, *** for p≤ 0.001

| Primary pulmonary hypertension |  |  |  |  | Secondary pulmonary hypertension |  |  |  |  |
| --- | --- | --- | --- | --- | --- | --- | --- | --- | --- |
|  | Cut-off | OR (95% CI) | Z value | P-value |  | Cut-off | OR (95% CI) | Z value | P-value |
| Score | 11.75 | 1.45 (1.14-2.16) | 5.76 | <0.0001*** | Score | 13.23 | 1.34 (1.2-2.01) | 18.11 | <0.0001*** |
| Score≥12.15 | - | 15.32 (10.34-27.45) | 11.87 | <0.0001*** | Score≥13.23 | - | 19.23 (11.88-37.21) | 24.13 | <0.0001*** |

### Machine learning results

The performance of FM model was compared to that of multivariate logistic regression, SVM, random forests, and multilayer perceptron, using significant univariable characteristics (without medications) as input to avoid overfitting (**Figure 1**). All of the models were trained with a randomly selected 80% (n=2048) of patients and tested with five-fold cross-validation approach using the remaining 20% (n=512) patients. The comparative performance evaluation results with metrics of recall, precision, F1-score and AUC were reported in **Table 7**.

**Table 7.** Performance comparison of Logistic Regression (LR), Random Forest (RF), Support Vector Machine (SVM), Multilayer Perceptron (MLP) in predicting primary/secondary pulmonary hypertension mortality risk with five-fold cross-validation approach (without medication predictors). The best metrics are underlined.

| Primary pulmonary hypertension |  |  |  |  | Secondary pulmonary hypertension |  |  |  |
| --- | --- | --- | --- | --- | --- | --- | --- | --- |
| Model | Precision | Recall | F1-score | AUC | Precision | Recall | F1-score | AUC |
| FM | <u>0.8966</u> | <u>0.8876</u> | <u>0.9106</u> | <u>0.9093</u> | <u>0.8693</u> | <u>0.8564</u> | <u>0.8864</u> | <u>0.8887</u> |
| LR | 0.7223 | 0.7614 | 0.7322 | 0.6659 | 0.7270 | 0.7306 | 0.7122 | 0.6770 |
| RF | 0.7619 | 0.7873 | 0.8000 | 0.8294 | 0.8233 | 0.8113 | 0.7876 | 0.8563 |
| SVM | 0.7838 | 0.8311 | 0.8234 | 0.8499 | 0.8323 | 0.8567 | 0.7973 | 0.7987 |
| MLP | 0.8258 | 0.8245 | 0.8514 | 0.8192 | 0.8330 | 0.8088 | 0.7964 | 0.8405 |

In the cross-validation, the FM model demonstrates significant improvement in pulmonary hypertension mortality risk prediction compared to other baselines. We can see that FM model outperforms baseline models with superior improvements for mortality prediction amongst both primary (precision=0.8966, recall=0.8876, F1-score=0.9106, AUC=0.9093) and secondary pulmonary hypertension patients (precision= 0.8693, recall= 0.8564, F1-score= 0.8864, AUC= 0.8887). As the important hyperparameters in the baseline models to improve prediction performance, for the SVM model the radial kernel parameters gamma and cost of constraints violation were tuned to 0.01, and 11, respectively. The number of trees and tree depth were tuned to be 732 and 9, respectively, for the random forest model. For the multilayer perceptron model, the number of units in the hidden layer was set to 5, and the decay was set to 0.073. The hyperparameter settings are finished with widely used grid and randomized search approach ^15^. The observations about model performance are consistent with previous studies that FM produced the most accurate predictions when compared to other models ^14, 16^.

## Discussion

In this study, we developed a predictive risk model for all-cause mortality in primary and secondary pulmonary hypertension patients incorporating baseline demographics, healthcare utilization metrics, comorbidities and drug prescription records using a population-based administrative database. A FM model was introduced as a multi-parametric mortality risk evaluation approach, which significantly improved risk prediction when compared to several baseline models.

The use of administrative databases for data mining in healthcare research has been a recent focus over the last decades. Specific to pulmonary hypertension, existing predictive risk models have largely relied on registry data. Whilst this can provide important insights by incorporating clinical parameters, such an approach can be difficult in the case of large patient numbers. In our study, we examined a territory-wide study of patients with both primary and secondary pulmonary hypertension, and used a multi-parametric approach incorporating data from different domains. We found common predictors of all-cause mortality for both primary and secondary causes, despite important differences in their aetiology, physiological basis and disease life course. The implications are that even without specific haemodynamic or physiological data, accurate predictions can be made.

### Pharmacotherapy and association with all-cause mortality

Regarding pharmacological treatment, diuretics are used in secondary pulmonary hypertension to reduce afterload ^17^. This is supported by their use to lower pulmonary arterial pressure in patients with right heart failure ^18^. In our study, the use of loop diuretics was significantly associated with a lower all-cause mortality. Moreover, whilst the efficacy of cardiac glycosides has not been extensively studied in pulmonary hypertension, one study found that the short-term use of digoxin can increase cardiac output for pulmonary hypertension patients with right ventricular failure ^19^ but not mortality ^20^. However, in contrary, our study found that their use was associated with higher mortality, which may be attributed to greater disease severity for patients who are prescribed these medications. Interestingly, the use of anti-arrhythmic drugs was also associated with higher mortality. This may suggest that patients with pulmonary hypertension die from causes other than cardiac arrhythmias. Unlike other drug groups in this study, the use of vasodilator antihypertensive drugs was not statistically significant in the prediction of all-cause mortality. A study found that patient response, i.e. pulmonary vasodilation, to the use of different vasodilators was highly variable between individuals ^21^. Furthermore, vasodilators such as calcium channel blockers are not considered empiric treatment for pulmonary hypertension and may only be effective as long-term treatment for a minority of patients who demonstrate an acute vasodilatory response ^22^. The highly individualized responsiveness and efficacy of vasodilators are hence a likely explanation as to why vasodilator antihypertensives were not a predictor of mortality.

### Factorization machine model for risk prediction

FM ^14^ as an efficient machine learning model has the main advantage stemming from its generality: a generic classifier working with any real-valued variable vector for supervised learning, in contract to matrix factorization ^23^ that only models the relation of two entities and over traditional machine learning models such as SVM ^24^, random forests ^25^, multilayer perceptron ^26^ that are quite difficult to capture the hidden interactions among characteristics in latent space. The main reason is that FM can learn meaningful embedding vectors for each variable as long as the variable itself appears enough times in the data, allowing the dot product a good estimator of pair-wise interaction effects even if two variables never or seldom co-occur. Previous experimental results demonstrated the better discrimination superiority of FM model in significantly improving prediction accuracy to be applied for multi-parametric mortality risk stratification in clinical practice.

### Strengths and limitations

The main strength of this study is the inclusion of a cohort of patients with pulmonary hypertension over an 18-year period with comprehensive laboratory, comorbidity and drug, healthcare utilization and follow-up data. This was complemented by machine learning analysis using characteristics of demographics, hospitalization, comorbidities and drug prescription data. Important informative indicators to predict mortality risk are detected using FM model, which demonstrate superior predictive performance over several baseline models.

However, several limitations should be noted. Firstly, long-term pulmonary hypertension mortality related comorbidity and disease onset sequence patterns are not uncovered. Secondly, clinical parameters from echocardiography, 6-minute walk tests and other physiological tests were not available in the administrative database and these variables could not be incorporated into our predictive risk models. These remain our future investigations to be explored.

## Conclusion

A machine learning model incorporating multi-modality clinical data significantly improves risk stratification performance and identify important indicators in predicting mortality in pulmonary hypertension.

## Data Availability

Data available upon request.

